# Wastewater surveillance for infectious disease: a systematic review

**DOI:** 10.1101/2021.07.26.21261155

**Authors:** Pruthvi Kilaru, Dustin Hill, Kathryn Anderson, Mary B. Collins, Hyatt Green, Brittany L. Kmush, David A. Larsen

**Author notes:** Correspondence to: Dr. David Larsen, Department of Public Health, Syracuse University, Syracuse, NY, 13244, USA.

## Abstract

Wastewater surveillance of SARS-CoV-2 has shown to be a valuable source of information regarding SARS-CoV-2 transmission and COVID-19 cases. Though the method has been used for several decades to track other infectious diseases, there has not been a comprehensive review outlining all of the pathogens surveilled through wastewater. The aim of this study is to identify what infectious diseases have been previously studied via wastewater surveillance prior to the COVID-19 Pandemic and identify common characteristics between the studies, as well as identify current gaps in knowledge. Peer-reviewed articles published as of August 1, 2020 that examined wastewater for communicable and infectious human pathogens on 2 or more occasions were included in the study. Excluded from this list were all reviews and methods papers, single collection studies, and non-human pathogens. Infectious diseases and pathogens were identified in studies of wastewater surveillance, as well as themes of how wastewater surveillance and other measures of disease transmission were linked. This review did not include any numerical data from individual studies and thus no statistical analysis was done. 1005 articles were identified but only 100 were included in this review after applying the inclusion criteria. These studies came from 38 countries with concentration in certain countries including Italy, Israel, Brazil, Japan, and China. Twenty-five separate pathogen families were identified in the included studies, with the majority of studies examining pathogens from the family Picornaviridae, including polio and non-polio enteroviruses. Most studies of wastewater surveillance did not link what was found in the wastewater to other measures of disease transmission. Among those studies that did compare wastewater surveillance to other measures of disease transmission the value observed was dependent upon pathogen and varied by study. Wastewater surveillance has historically been used to assess water-borne and fecal-orally transmitted pathogens causing diarrheal disease. However, numerous other types of pathogens have been surveilled using wastewater and wastewater surveillance should be considered as a potential tool for many infectious diseases. Wastewater surveillance studies can be improved by incorporating other measures of disease transmission at the population-level including disease incidence and hospitalizations.

## Introduction

Infectious disease surveillance is most commonly conducted at the health center or the hospital,^1^ either through passive reporting or active case finding.^2^ This type of event-based infectious disease surveillance monitors trends in morbidity and mortality, alerting health systems when a statistically improbable uptick of events occurs. In this way, the number of cases, hospitalizations, and deaths from endemic infectious diseases such as malaria or influenza are tracked and the effectiveness of interventions such as mosquito control or vaccines can be monitored. Importantly, due to cost and non-representative access to molecular diagnostics, many pathogens under surveillance are characterized by their symptoms or syndromes, such as influenza-like illness. For emerging pathogens, event-based infectious disease surveillance may note an odd increase in some symptom or condition, notably as occurred with microcephaly and Zika,^3,4^ or pneumonia cases without a known cause as occurred with COVID-19.^5^ Event-based infectious disease surveillance requires a health system capable of observing an unexpected trend, a population with sufficient access to that health system, and a sufficiently large trend or cluster of odd cases to alert officials.

Environmental surveillance, on the other hand, is a broad category for systems that monitor the presence or absence of a pathogen in the environment. Their defining characteristic is the circumvention of human behavior and health systems, which reduces bias, while still providing information regarding risks to human health. For example, environmental surveillance may routinely test known vectors for pathogens,^6^ alerting the public to the detection of, or an increase in, the pathogen in the vector population.

Wastewater surveillance is a type of environmental surveillance that has historically been utilized to track water-borne or fecal-orally transmitted pathogens. The origins of wastewater surveillance hail back to the London cholera epidemic of the mid-1800’s, John Snow, and the Broad Street Pump, when a cesspool near a house with multiple cholera deaths was excavated and found to be leaking into the pump’s water supply.^7^ With the scientific evidence supporting germ theory, scientists began hunting sewage not only for cholera but also for other pathogens including salmonella typhi bacteria (typhoid),^8,9^ coxsackie viruses,^10^ and poliovirus.^11^ From the 1970’s onward, wastewater surveillance formed a critical component of the worldwide initiative to eradicate polio,^12^ and perhaps polio provides the best contrast between event-based and environmental surveillance systems. Whereas event-based polio surveillance relies on an unexpected increase in acute flaccid paralysis which occurs in only 0.5% of polio cases,^13^ wastewater surveillance can detect poliovirus circulating in a community before any paralysis occurs.^14^

The COVID-19 pandemic saw the broad adaptation of wastewater surveillance across the globe,^15^ as the limitations of event-based surveillance systems for an emerging pathogen were laid bare. Most interestingly, COVID-19 is a respiratory-transmitted pathogen, suggesting that a pathogen’s mode of transmission need not be fecal-oral or waterborne for wastewater surveillance to be useful. Could wastewater surveillance be a more widely applied tool, not only monitoring trends in waterborne or fecal-orally transmitted pathogens but also pathogens of pandemic potential and those causing the greatest burden of disease? SARS-CoV-2 is certainly the pathogen du jour, but as wastewater surveillance systems are erected should they also be incorporating other pathogens into their surveillance? Herein we present a systematic review of wastewater surveillance for infectious disease, reporting the documented successes of testing wastewater for infectious disease pathogens that circulate primarily in humans.

## Methods

### Systematic Literature Review

Following PRISMA guidelines^16^, we searched PubMed, SCOPUS, Science Direct and Google Scholar for studies looking at wastewater-based surveillance of infectious diseases (both viral and bacterial) in human populations and published before August 1st, 2020. For the databases (PubMed, SCOPUS, and Science Direct), search terms included Mesh headings, MeSH terms, and text words and synonyms, including “Wastewater”, “Waste water”, “Sewage”, “Sewer”, “Environmental”, “Surveillance”, “Disease”, “Feces”, “wastewater-based epidemiology”, “Environmental surveillance”, “Environmental Epidemiology “, “Wastewater Surveillance “, “Environmental Monitoring”, “Wastewater Monitoring”, “Virus”, “Bacteria”. These terms were combined using the boolean terms “AND” and “OR” when applicable. Similar terms were used but with filters on Google Scholar to limit the search to material of interest. The filters included the inclusion of the characters “doi” to look for a Digital Object Identifier to ensure that it was a published work, and the exclusion of the terms “systematic review”, “literature review”, “meta-analysis”, and “review” in the title. The boolean term “NOT” was used to aid in excluding these terms. All sources, databases and Google Scholar, were filtered to look for texts in the English language. The search string used for each individual database and Google Scholar, as well as the filters used, can be found in Appendix 1.

Once article lists were pulled from their respective sources, duplicates were removed, using Microsoft Excel’s built-in remove duplicated function, using both title and authors as the reference for removal. Reviewers (Pruthvi Kilaru and Dustin Hill) screened titles and abstracts for remaining articles, retrieved articles for full-text review, and assessed full-text articles based on eligibility criteria.

### Eligibility Criteria

We included published studies which tested wastewater for communicable and/or infectious human diseases on more than one occasion and during two or more time periods. Non-communicable diseases, such as diabetes and obesity, were excluded. As we defined surveillance as having the requirement of testing over time, all articles which tested wastewater only once and/or on a single day were excluded. Articles which discussed diseases not related to humans or not in the context of humans (e.g. influenza virus in pigs), were also excluded. Peer-reviewed journal articles were included as long as they were not reviews, systematic reviews, literature reviews, or meta-analyses. Non-peer reviewed journal articles such as research notes, research letters, and short communications were excluded. Methods papers that looked purely at and compared different techniques of drawing and sampling wastewater were also excluded if they did not offer analysis of pathogens naturally present in the wastewater. This included studies that spike wastewater with a pathogen only to look at recovery in the context of comparing methods of sampling. Lastly, we excluded all papers which reported the surveillance of SARS-CoV0-2. This determination was made to support the utility of environmental surveillance outside of emergency/pandemic situations, to determine what and where disease surveillance has been conducted in the past, and to support expansion and extension of surveillance to other pathogens and regions.

### Data Extraction

We initially extracted the following information from the articles meeting the eligibility criteria: period of sampling, country the sampling occurred in, pathogen(s)/disease(s) being monitored, number of samples pulled, amount of sample pulled, sample type (grab, composite, other), method of detection, overall findings, was genetic typing done, and did the researchers connect their findings to population health. The primary information of interest were the disease(s) being monitored, method of detection, and if the authors connected their findings to population health.

## Results

Literature searches initially identified 1005 entries (after removing duplicates), of which 159 abstracts met the inclusion criteria. After review of the articles, 100 scientific papers were included (Figure 1, Table 1).

**Figure 1.**
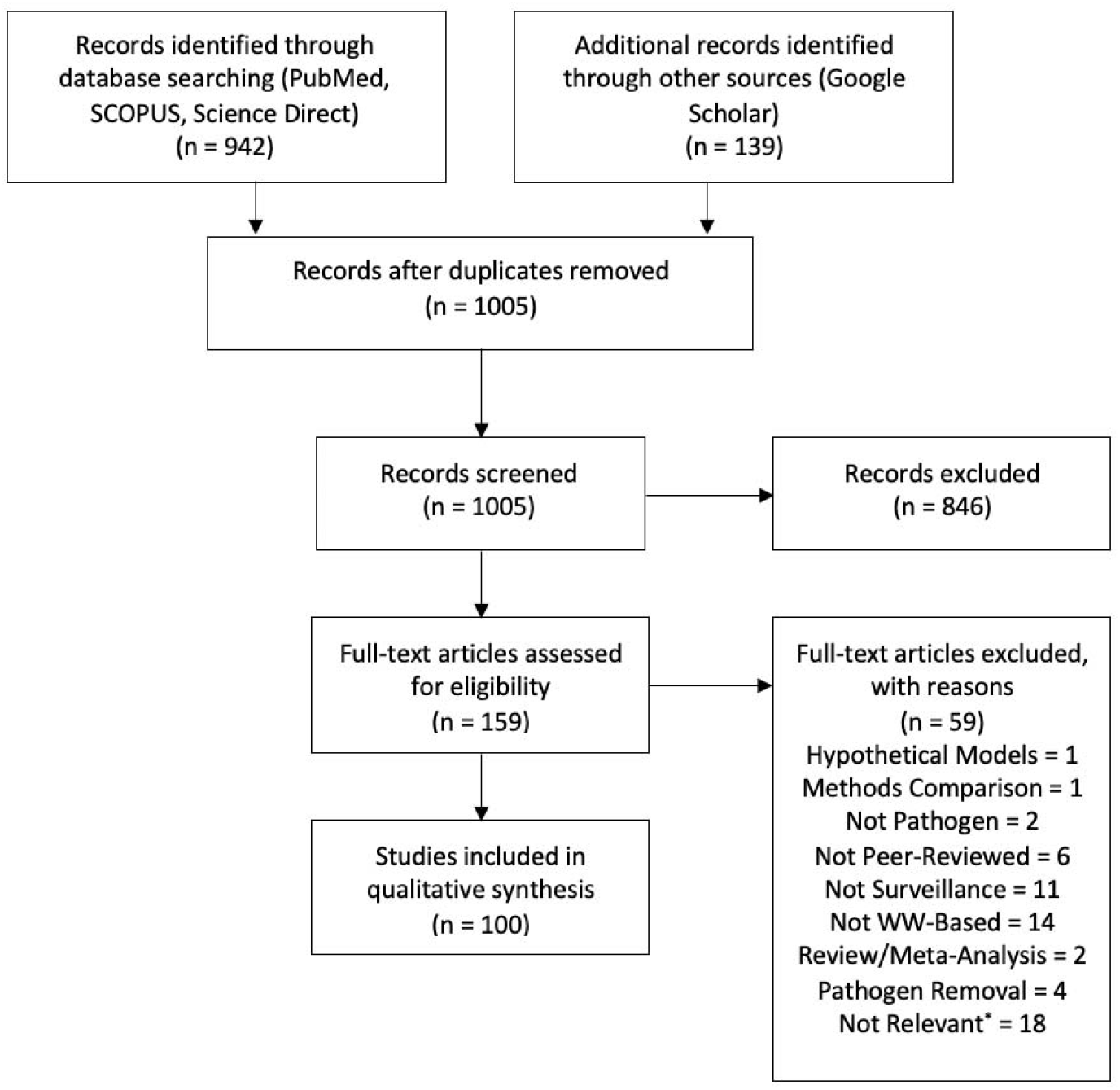
PRISMA flow chart of articles included in the review. Reasons for exclusion include: hypothetical Models – the experiment was hypothetical and no data were collected; methods comparison – the paper compared multiple recovery methods; not pathogen – paper focused on non-communicable diseases (e.g. diabetes); not surveillance – sampled only once or for non-surveillance purposes; not WW-based – wastewater was not directly tested; pathogen removal – paper looked at removal techniques of pathogens in wastewater; not relevant* - e.g. diseases not tied to human population, effect on other species/animals

**Table 1:**
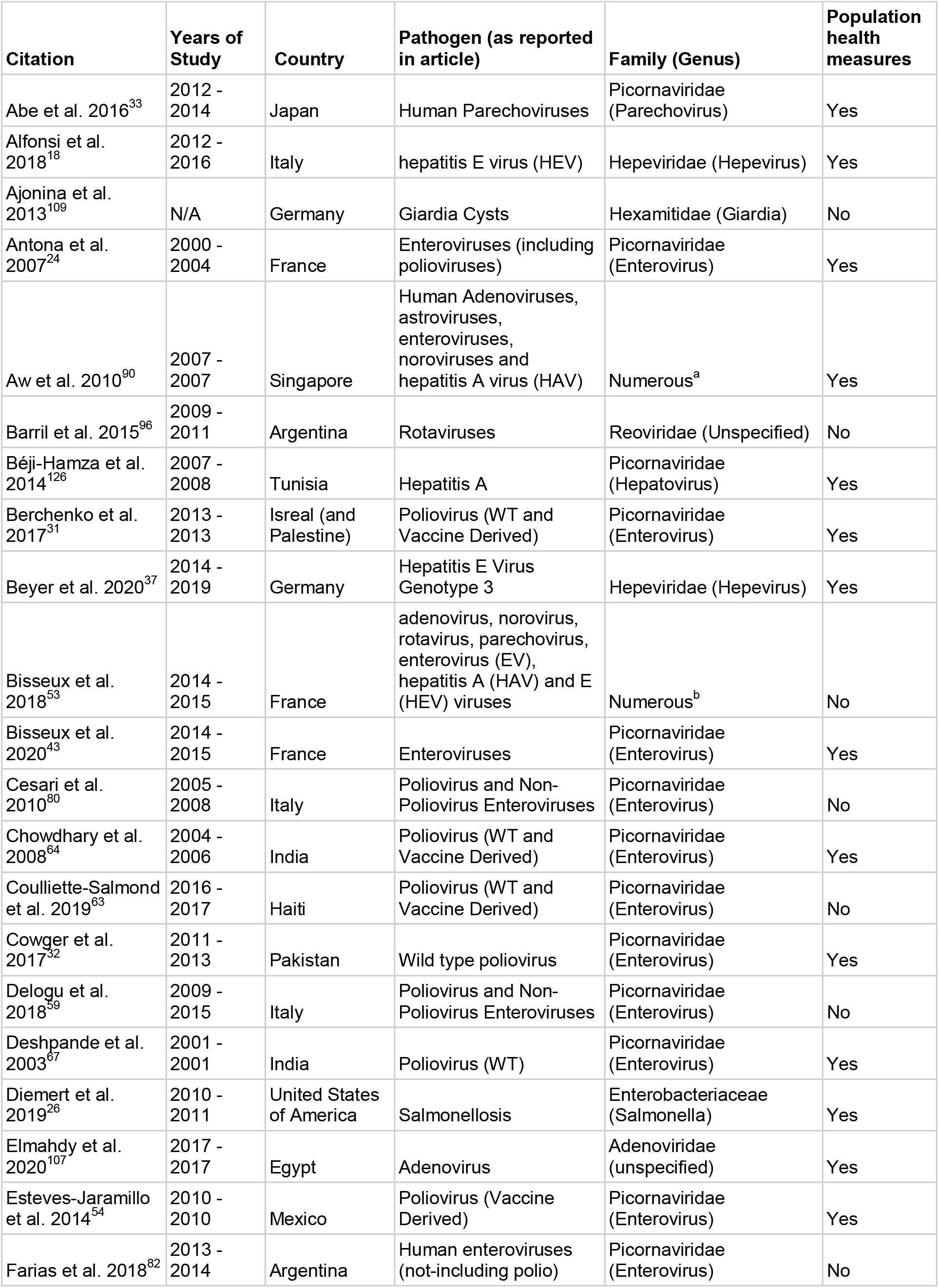
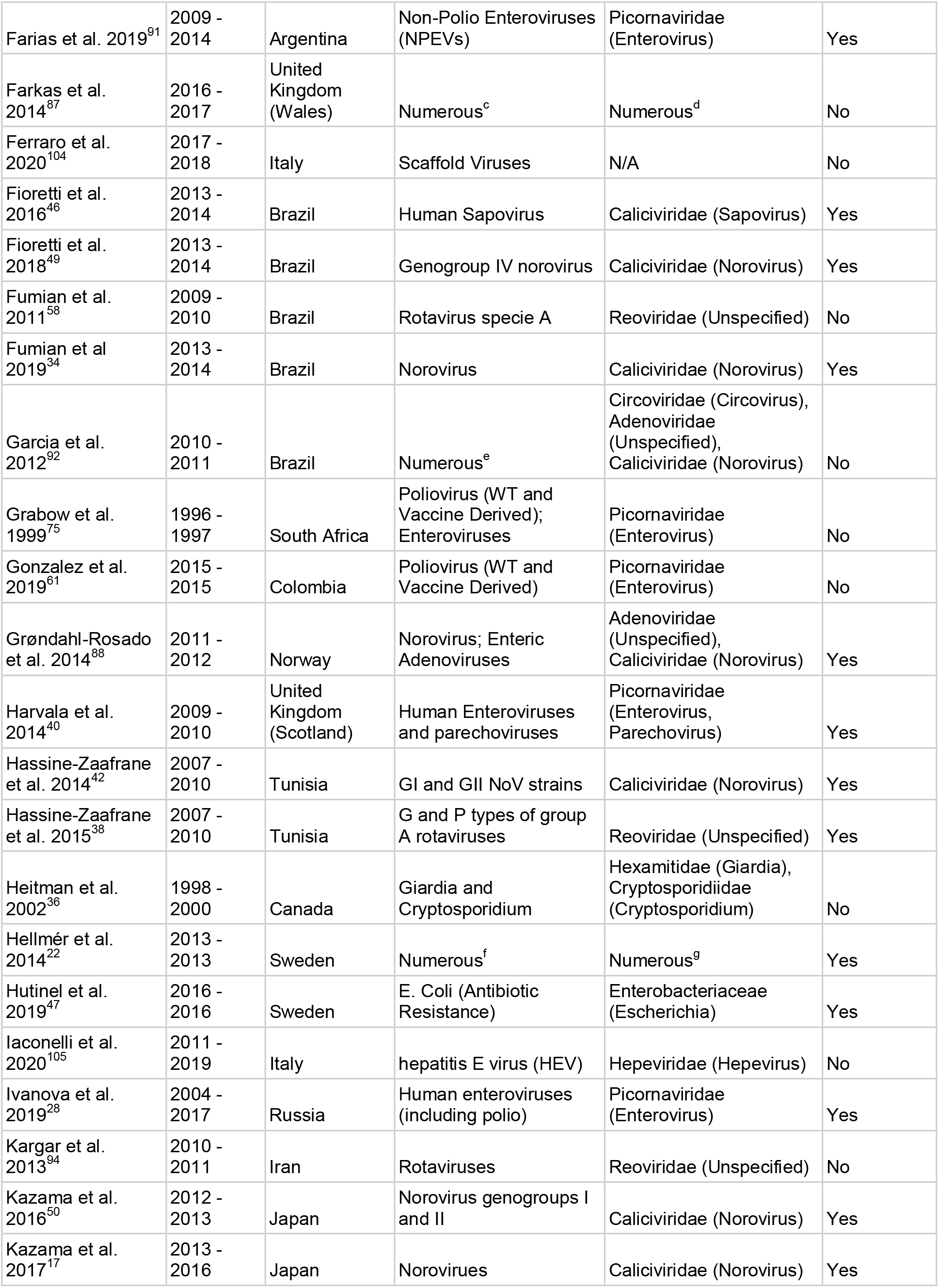
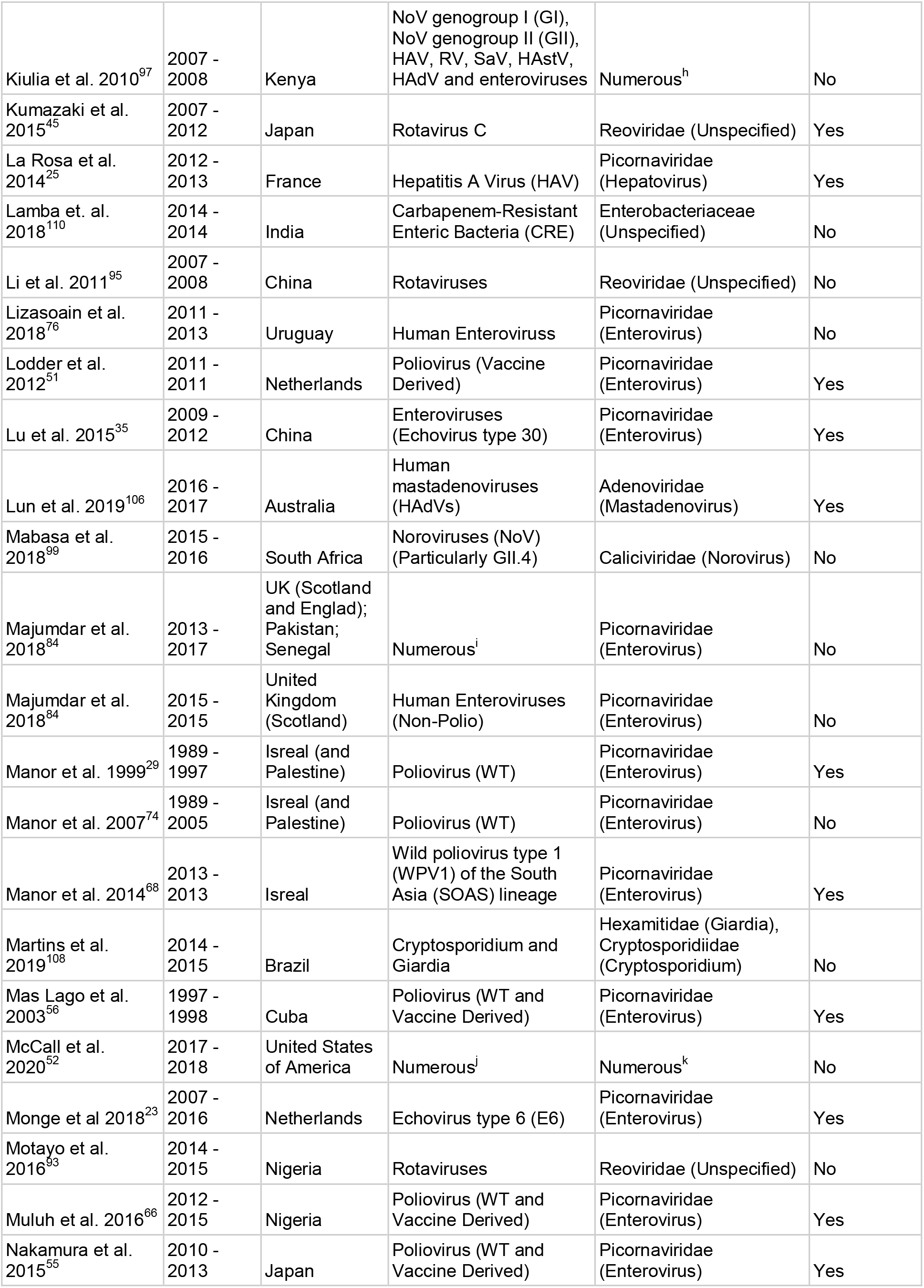
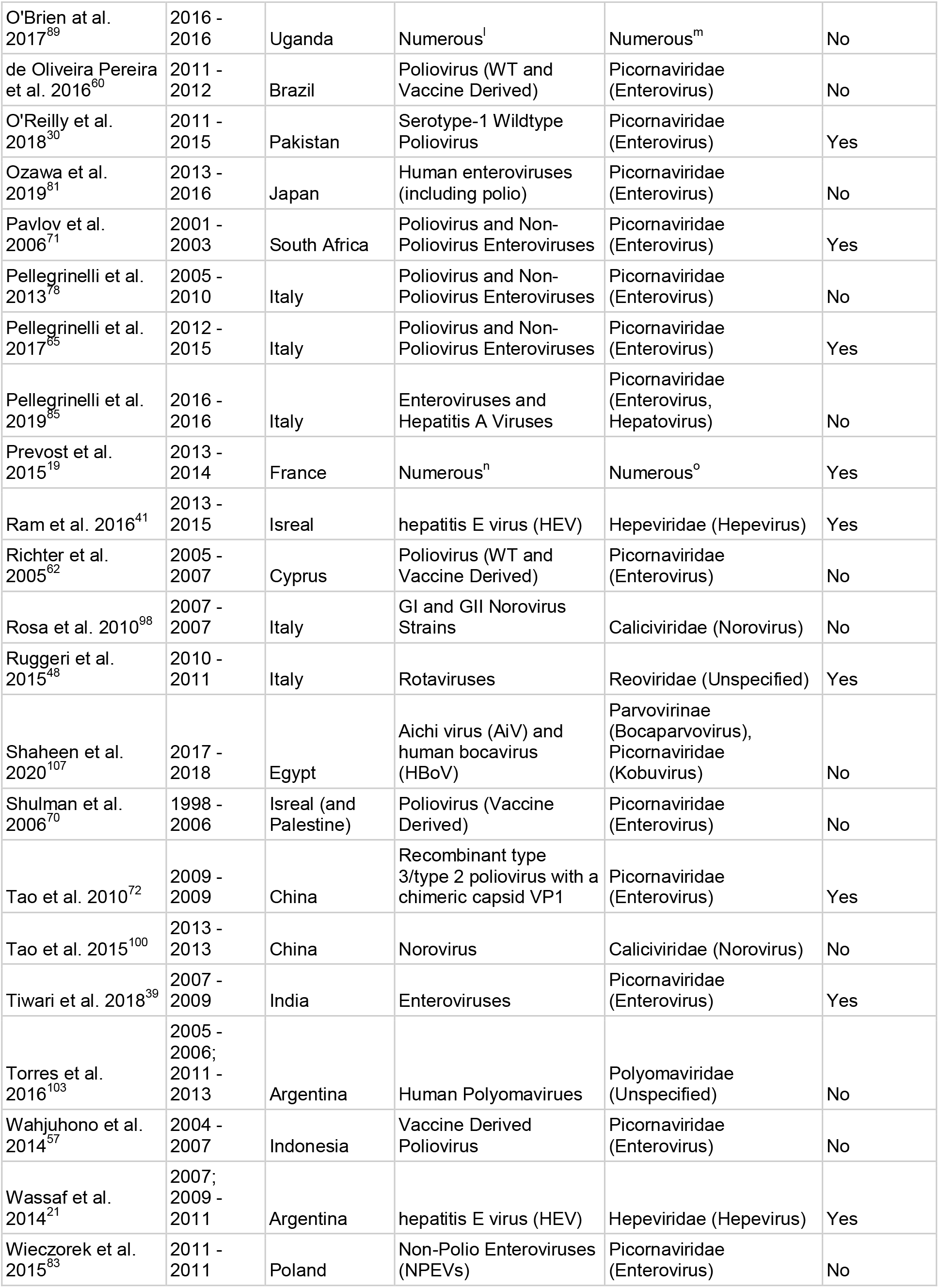
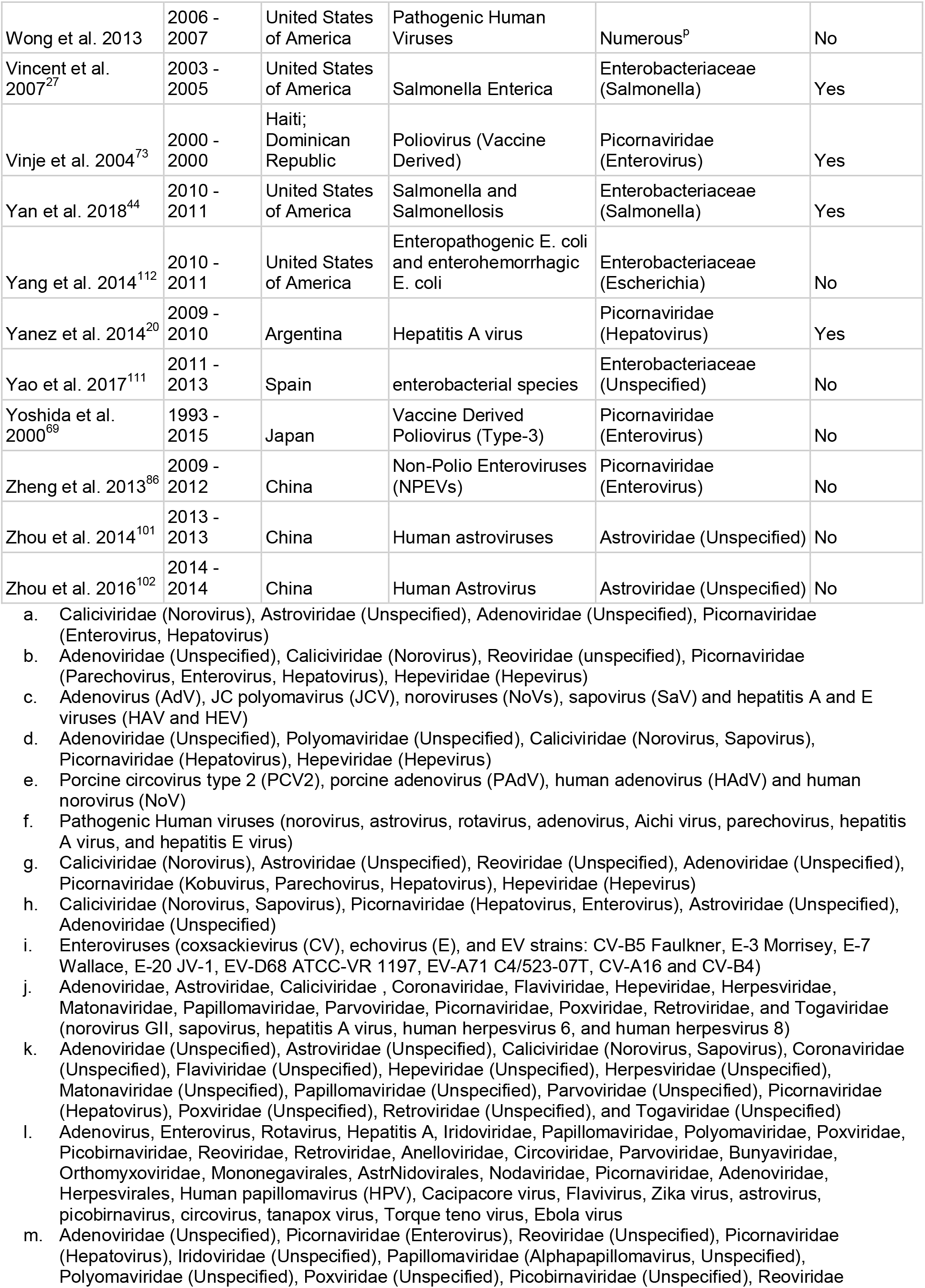
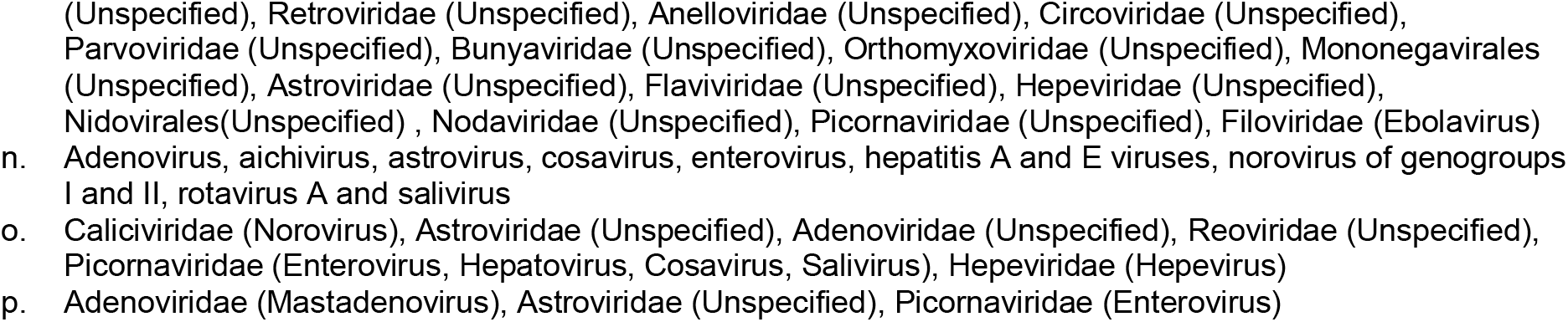
Characteristics of wastewater surveillance studies included in the systematic review.

Across the 100 included articles, studies were conducted in 38 countries with the most studies conducted in Italy (10 studies), China (8 studies), Japan (7 studies), Israel (7 studies), and Brazil (7 studies). These 5 countries accounted for 39% (39/100) of the studies conducted across all articles.

Within the included articles, the most prevalent pathogens found were viruses from the families Picornaviridae, Calciviridae, Adenoviridae, Reoviridae, and Hepeviridae (Figure 3). Of the most prevalent families, three of them are known to have pathogens contributing to diarrheal diseases (Picornaviridae, Caliciviridae, and Reoviridae) and make up 57.5% of the pathogens studied across all articles. Within the Piconaviridae family, the most prevalent genus studied was enteroviruses, with poliovirus being the most popular among that genus. Enteroviruses made up 32.5% (52 instances) of pathogens found in all of the articles. Additionally, there were 20 other families of pathogens that appeared between 1 - 9 times within our literature review, with a mean of 2.2 appearances and a median of 2 appearances each. Considering the global burden of disease (Figure 4), diarrheal diseases were the most represented among studies of wastewater surveillance, with other infectious diseases with a great burden not found in this systematic review. Infectious diseases of international concern were better represented, with only influenza and HIV/AIDS not represented among studies of wastewater surveillance (Figure 4).

**Figure 2.**
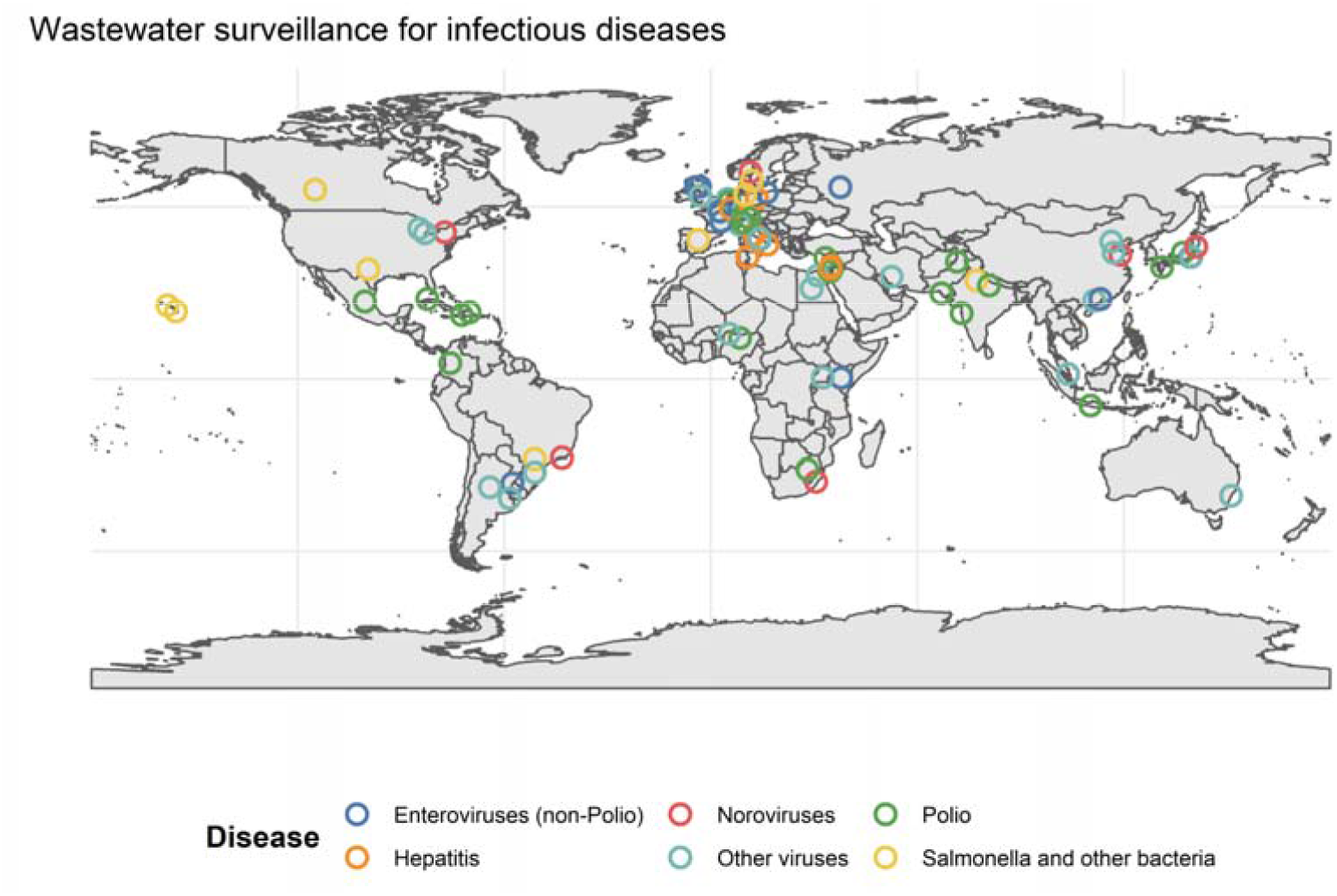
Global distribution of studies of wastewater surveillance for infectious disease.

**Figure 3.**
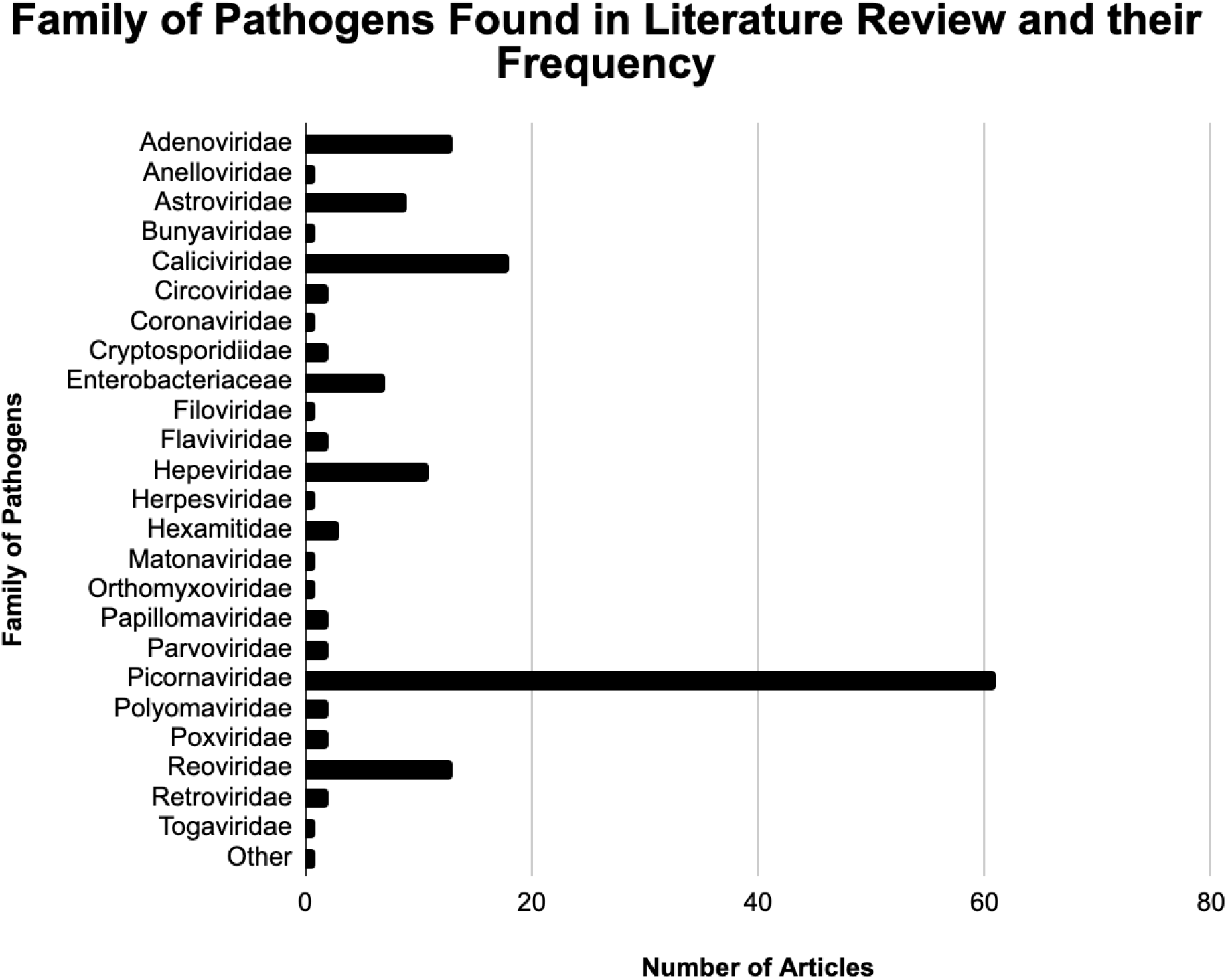
Bar graph showing the families of pathogens found in the included articles and their frequency.

**Figure 4.**
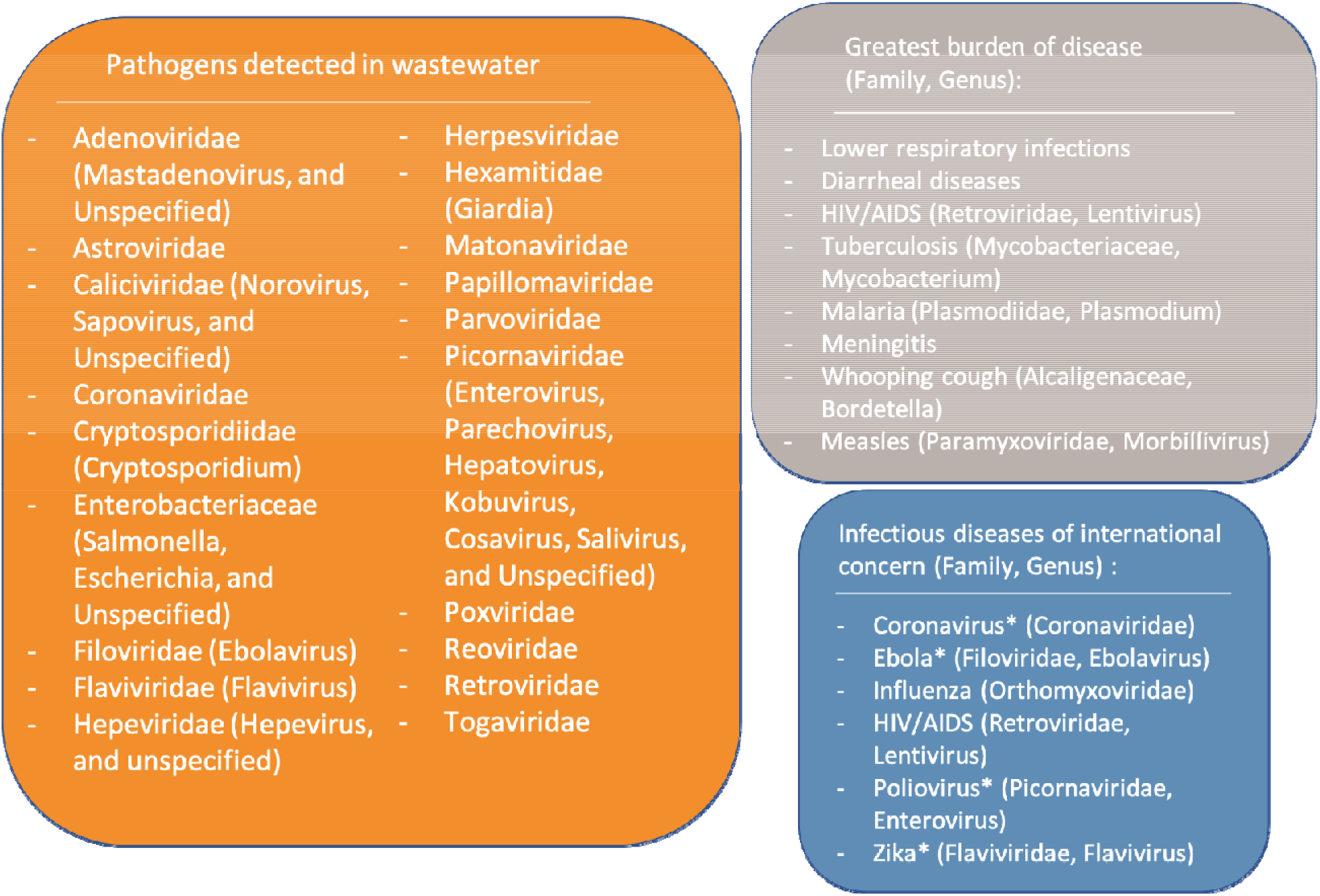
Pathogens surveilled in wastewater (orange group) are not reflected in the greatest burden of disease except for diarrheal diseases (gray group). Many infectious diseases of international concern (blue group) have been surveilled in wastewater, however as represented by an asterisk (*).

A number of studies correlated the level of an infectious disease pathogen found in wastewater to relevant measures of transmission such as population-level incidence, without reporting if public health action or policy was influenced by wastewater surveillance or not. Studies have linked the level of norovirus in wastewater to incidence of gastroenteritis,^17^ levels of hepatitis E virus in wastewater to incidence of hepatitis E,^18^ and level of enteric viruses in wastewater to the incidence of acute diarrhea.^19^ Other studies compared population seroprevalence to the level of hepatitis A virus^20^ and hepatitis E virus^21^ found in wastewater. In comparison with the incidence of clinical cases, wastewater surveillance provided early warning of hepatitis A virus and norovirus outbreaks in Sweden.^22^ However, in the Netherlands wastewater surveillance did not serve well in an early warning capacity for a variety of enteroviruses.^23^ Wastewater surveillance correlated well with outbreaks of enterovirus,^24^ hepatitis A virus,^25^ and *Salmonella enterica*.^26,27^ In Russia, outbreaks of aseptic meningitis caused by echovirus type 6 correlated with levels in wastewater, but outbreaks of aseptic meningitis caused by echovirus type 30 did not.^28^ A few studies directly compared the sensitivity of surveillance for the incidence of acute flaccid paralysis (a type of non-specific clinical surveillance) to wastewater surveillance for poliovirus, finding that wastewater surveillance was more sensitive and combining the two systems was optimal.^29–32^ The most common type of comparison of wastewater surveillance with clinical cases linked the genetic diversity of bacteria or viral strains found in wastewater surveillance back to samples from clinical cases of meningitis, gastroenteritis, or diarrhea-related illness.^33–53^ These studies did not examine the use of wastewater surveillance to inform of outbreaks or correlate levels of the pathogens found in wastewater to trends in population-level incidence over time.

A handful of publications documented the utility of wastewater surveillance to assess the impact of public health interventions. Wastewater surveillance was able to confirm the cessation of the transmission of vaccine-derived poliovirus following a transition from oral poliovirus vaccine to inactivated poliovirus vaccine in numerous studies.^54–57^ Wastewater surveillance was also used to assess the impact of rotavirus vaccine deployment in Rio de Janeiro, Brazil.^58^

When considering the use of wastewater surveillance to inform public health action or policy, the most common reported application was to document the elimination of wildtype poliovirus transmission.^59–65^ In countries with circulation of wildtype poliovirus, wastewater surveillance has been used to guide vaccination efforts. In Nigeria, directed vaccine efforts based on results from wastewater surveillance interrupted polio transmission in numerous areas.^66^ In Mumbai, India, wastewater surveillance was used to alert importation of wild-type poliovirus and inform subsequent vaccine distributions.^67^ And in Israel, the importation of wildtype poliovirus was detected using wastewater surveillance which then led to an expansion of wastewater surveillance and vaccination campaigns to prevent re-establishment of poliovirus transmission.^68^ No articles were identified that documented the use of wastewater surveillance to inform public health action for any other pathogen than poliovirus.

The majority of articles reporting on wastewater surveillance included no comparison to other measures of transmission such as clinical cases of disease. Some articles assessed the presence of poliovirus,^69–72^ either wildtype or vaccine-derived, including potential neurovirulence of vaccine-derived poliovirus.^73–75^ Many articles documented the diversity of non-polio enteroviruses found in wastewater, ^73,76–92^ with a variety of focuses including rotavirus,^93–97^ norovirus,^88,92,98–100^ astrovirus,^101,102^ polyomavirus,^103^ Saffold virus,^104^ hepatitis A virus,^85^ hepatitis E virus,^105^ mastadenovirus,^106^ Aichi virus,^107^ and human bocavirus.^107^ Surveillance of *Giardia* and/or *Cryptosporidium* was also documented.^108,109^ Other studies examined the extent of antimicrobial resistance,^110,111^ or virulence genes,^112^ in *Escherichia coli* or *Salmonella* bacteria.

## Discussion

We found that wastewater surveillance has been used extensively to guide public health policy and interventions to eliminate and eradicate poliovirus, but we found no reports of wastewater surveillance being used proactively for other pathogens. With the COVID-19 pandemic, wastewater surveillance has been proactively used by a variety of organizations, including institutions of higher education,^113^ local health departments, and national governments. Linking wastewater surveillance to public health interventions, however, can be challenging. From our review, the most obvious link between wastewater surveillance and public health policy/intervention was the confirmation of the absence of transmission of polio, as well as early notification or confirmation of outbreaks. Linking wastewater surveillance to population-level incidence should also be straightforward, but we found relatively few studies doing so. There certainly is difficulty in obtaining incidence rates for a variety of pathogens, but this should not prevent scientists from comparing wastewater surveillance to syndromic surveillance, e.g. incidence of diarrhea, gastroenteritis, or pneumonia. Increased collaboration between epidemiologists, microbiologists, and environmental engineers is needed to maximize the knowledge gained from studies of wastewater surveillance.

As evidenced in this review, epidemiologists have typically thought of wastewater surveillance only as a tool to surveil pathogens that are either waterborne or fecal-orally transmitted. For example, a recent textbook highlights the potential for wastewater surveillance for waterborne pathogens, but completely ignores pathogens of other transmission types.^114^ This narrow focus overlooks the potential utility of wastewater surveillance for sexually-transmitted, respiratory-transmitted, and vector-borne diseases of pandemic potential.^115,116^ Indeed, only one of the six times that the World Health Organization has declared a public health emergency of international concern (a term conceptualized in 2005) has the pathogen been waterborne or fecal-orally transmitted (poliovirus compared to H1N1 influenza, ebola twice, Zika, and COVID-19).^117^ In addition, the only pandemics in the 20th century were caused by influenza and HIV/AIDS.

The COVID-19 pandemic has shown wastewater surveillance to be an effective tool for a respiratory-transmitted pathogen.^118^ Given the low cost and population-level representation that a single wastewater sample provides, further research into the utility of wastewater surveillance for infectious diseases in general is needed. Among other pathogens that are not waterborne nor fecal-orally transmitted, we found reports of Zika and Ebola virus in wastewater, suggesting that they could be potential targets of continuous wastewater surveillance. Wastewater surveillance could be useful for other high-burden infectious diseases as well. Evidence from the 1990’s suggests HIV can be detected in wastewater,^119^ although this systematic review found no reports of surveilling HIV in wastewater. Tuberculosis can also be found in wastewater,^120^ even to the extent of endangering sewage workers.^121^ But again this systematic review found no reports of surveilling tuberculosis in wastewater. Bearing in mind that wastewater surveillance is useful for tracking antimicrobial resistance^122^ should wastewater be useful for surveilling tuberculosis, then it could potentially be used to surveil multi-drug resistant tuberculosis as well. Malaria can be easily diagnosed in human feces,^123^ which leaves us to speculate the possibility for finding and surveilling this pathogen in wastewater. Numerous groups are currently assessing the capacity to find influenza in wastewater, but H1N1 influenza was not found in the wastewater of the Netherlands during the 2009 pandemic.^124^

Wastewater surveillance should be considered a general tool for public health going forward. In order to maximize its utility, further understanding of what pathogens can and cannot be surveilled using wastewater is needed. A variety of factors affect the probability that a pathogen will be found in wastewater and then documented in the scientific literature.

Publication bias certainly plays a role, with numerous pathogens neglected in the scientific literature,^125^ as well as negative results not being published. Along these lines different types of studies are needed including studies of load shedding dynamics, pathogens’ persistence in wastewater, and the relationship between levels of a pathogen found in wastewater and other measures of transmission such as population-level incidence. Perhaps most important for public health, more studies are needed that assess the utility of wastewater surveillance to guide policy and public health intervention.

## Data Availability

Data in the article is available from the authors upon reasonable request.

## Contributors

Conceptualization: DAL

Data curation: PK

Formal analysis: N/A

Funding acquisition: N/A

Investigation: PK, DH

Methodology: KA, MBC, HG, BLK, DAL

Project administration: DAL

Resources: N/A

Software: N/A

Supervision: DAL, MBC

Validation: N/A

Visualization: PK, DH, MBC, DAL

Writing - original draft: PK, DAL

Writing - reviewing and editing: PK, DH, KA, MBC, HG, BLK, DAL

## Data sharing

No data were collected as part of this study.

## Acknowledgements

None.

## References

1 Abat C, Chaudet H, Rolain J-M, Colson P, Raoult D. Traditional and syndromic surveillance of infectious diseases and pathogens. International Journal of Infectious Diseases 2016; 48: 22–8.

2 Murray J, Cohen AL. Infectious Disease Surveillance. International Encyclopedia of Public Health 2017; : 222–9.

3 de Araújo TVB, Rodrigues LC, de Alencar Ximenes RA, et al. Association between Zika virus infection and microcephaly in Brazil, January to May, 2016: preliminary report of a case-control study. The Lancet Infectious Diseases 2016; 16: 1356–63.

4 Schuler-Faccini L, Ribeiro EM, Feitosa IML, et al. Possible association between Zika virus infection and microcephaly — Brazil, 2015. Morbidity and Mortality Weekly Report 2016; 65: 59–62.

5 Zhu N, Zhang D, Wang W, et al. A Novel Coronavirus from Patients with Pneumonia in China, 2019. New England Journal of Medicine 2020; 382: 727–33.

6 Gu W, Unnasch TR, Katholi CR, Lampman R, Novak RJ. Fundamental issues in mosquito surveillance for arboviral transmission. Transactions of The Royal Society of Tropical Medicine and Hygiene 2008; 102: 817–22.

7 Johnson S. The Ghost Map: The Story of London’s Most Terrifying Epidemic--and how it Changed Science, Cities, and the Modern World. Penguin, 2006.

8 Moore B. The detection of enteric carriers in towns by means of sewage examination. J R Sanit Inst 1951; 71: 57–60.

9 Gray JDA. The isolation of B. Paratyphosus B from sewage. Br Med J 1929; 1: 142–4.

10 Kelly SM, Clark ME, Coleman MB. Demonstration of Infectious Agents in Sewage. Am J Public Health Nations Health 1955; 45: 1438–46.

11 Paul JR, Trask JD, Gard S. Ii. Poliomyelitic virus in urban sewage. Journal of Experimental Medicine 1940; 71: 765–77.

12 Asghar H, Diop OM, Weldegebriel G, et al. Environmental surveillance for polioviruses in the global polio eradication initiative. Journal of Infectious Diseases 2014; 210: S294–303.

13 Tangermann RH, Lamoureux C, Tallis G, Goel A. The critical role of acute flaccid paralysis surveillance in the Global Polio Eradication Initiative. International Health 2017; 9: 156–63.

14 Brouwer AF, Eisenberg JNS, Pomeroy CD, et al. Epidemiology of the silent polio outbreak in Rahat, Israel, based on modeling of environmental surveillance data. Proceedings of the National Academy of Sciences of the United States of America 2018; 115: E10625–33.

15 Naughton CC, Roman FA, Alvarado AGF, et al. Show us the Data: Global COVID-19 Wastewater Monitoring Efforts, Equity, and Gaps. Public and Global Health, 2021 DOI:10.1101/2021.03.14.21253564.

16 Moher D, Liberati A, Tetzlaff J, Altman DG. Preferred reporting items for systematic reviews and meta-analyses: the PRISMA statement. BMJ 2009; 339: b2535.

17 Kazama S, Miura T, Masago Y, et al. Environmental Surveillance of Norovirus Genogroups I and II for Sensitive Detection of Epidemic Variants. Appl Environ Microbiol 2017; 83. DOI:10.1128/AEM.03406-16.

18 Alfonsi V, Romanò L, Ciccaglione AR, et al. Hepatitis e in Italy: 5 years of national epidemiological, virological and environmental surveillance, 2012 to 2016. Eurosurveillance 2018; 23. DOI:10.2807/1560-7917.ES.2018.23.41.1700517.

19 Prevost B, Lucas FS, Goncalves A, Richard F, Moulin L, Wurtzer S. Large scale survey of enteric viruses in river and waste water underlines the health status of the local population. Environ Int 2015; 79: 42–50.

20 Yanez LA, Lucero NS, Barril PA, et al. Evidence of hepatitis A virus circulation in central Argentina: seroprevalence and environmental surveillance. J Clin Virol 2014; 59: 38–43.

21 Martínez Wassaf MG, Pisano MB, Barril PA, et al. First detection of hepatitis E virus in Central Argentina: environmental and serological survey. J Clin Virol 2014; 61: 334–9.

22 Hellmér M, Paxéus N, Magnius L, et al. Detection of Pathogenic Viruses in Sewage Provided Early Warnings of Hepatitis A Virus and Norovirus Outbreaks. Appl Environ Microbiol 2014; 80: 6771–81.

23 Monge S, Benschop K, Soetens L, et al. Echovirus type 6 transmission clusters and the role of environmental surveillance in early warning, the Netherlands, 2007 to 2016. Euro Surveill 2018; 23. DOI:10.2807/1560-7917.ES.2018.23.45.1800288.

24 Antona D, Lévêque N, Chomel JJ, Dubrou S, Lévy-Bruhl D, Lina B. Surveillance of enteroviruses in France, 2000-2004. Eur J Clin Microbiol Infect Dis 2007; 26: 403–12.

25 La Rosa G, Della Libera S, Iaconelli M, et al. Surveillance of hepatitis A virus in urban sewages and comparison with cases notified in the course of an outbreak, Italy 2013. BMC Infectious Diseases 2014; 14: 1–11.

26 Diemert S, Yan T. Clinically Unreported Salmonellosis Outbreak Detected via Comparative Genomic Analysis of Municipal Wastewater Salmonella Isolates. Appl Environ Microbiol 2019; 85. DOI:10.1128/AEM.00139-19.

27 Vincent V, Scott HM, Harvey RB, Alali WQ, Hume ME. Novel surveillance of Salmonella enterica serotype Heidelberg epidemics in a closed community. Foodborne Pathog Dis 2007; 4: 375–85.

28 Ivanova OE, Yarmolskaya MS, Eremeeva TP, et al. Environmental Surveillance for Poliovirus and Other Enteroviruses: Long-Term Experience in Moscow, Russian Federation, 2004–2017. Viruses 2019; 11. DOI:10.3390/v11050424.

29 Manor Y, Handsher R, Halmut T, et al. Detection of Poliovirus Circulation by Environmental Surveillance in the Absence of Clinical Cases in Israel and the Palestinian Authority. J Clin Microbiol 1999; 37: 1670–5.

30 O’Reilly KM, Verity R, Durry E, et al. Population sensitivity of acute flaccid paralysis and environmental surveillance for serotype 1 poliovirus in Pakistan: an observational study. BMC Infect Dis 2018; 18: 176.

31 Berchenko Y, Manor Y, Freedman LS, et al. Estimation of polio infection prevalence from environmental surveillance data. Sci Transl Med 2017; 9. DOI:10.1126/scitranslmed.aaf6786.

32 Cowger TL, Burns CC, Sharif S, et al. The role of supplementary environmental surveillance to complement acute flaccid paralysis surveillance for wild poliovirus in Pakistan - 2011-2013. PLoS One 2017; 12: e0180608.

33 Abe M, Ueki Y, Miura T, et al. Detection of Human Parechoviruses in Clinical and Municipal Wastewater Samples in Miyagi, Japan, in 2012–2014. Japanese Journal of Infectious Diseases 2016; 69: 414–7.

34 Fumian TM, Fioretti JM, Lun JH, dos Santos Ial, White PA, Miagostovich MP. Detection of norovirus epidemic genotypes in raw sewage using next generation sequencing. Environment International 2019; 123: 282–91.

35 Lu J, Zheng H, Guo X, et al. Elucidation of Echovirus 30’s Origin and Transmission during the 2012 Aseptic Meningitis Outbreak in Guangdong, China, through Continuing Environmental Surveillance. Appl Environ Microbiol 2015; 81: 2311–9.

36 Heitman TL, Frederick LM, Viste JR, et al. Prevalence of Giardia and Cryptosporidium and characterization of Cryptosporidium spp. isolated from wildlife, human, and agricultural sources in the North Saskatchewan River Basin in Alberta, Canada. Canadian Journal of Microbiology 2002. DOI:10.1139/w02-047.

37 Beyer S, Szewzyk R, Gnirss R, Johne R, Selinka H-C. Detection and Characterization of Hepatitis E Virus Genotype 3 in Wastewater and Urban Surface Waters in Germany. Food Environ Virol 2020; 12: 137–47.

38 Hassine-Zaafrane M, Kaplon J, Ben Salem I, et al. Detection and genotyping of group A rotaviruses isolated from sewage samples in Monastir, Tunisia between April 2007 and April 2010. J Appl Microbiol 2015; 119: 1443–53.

39 Tiwari S, Dhole TN. Assessment of enteroviruses from sewage water and clinical samples during eradication phase of polio in North India. Virology Journal 2018; 15: 157.

40 Harvala H, Calvert J, Nguyen DV, et al. Comparison of diagnostic clinical samples and environmental sampling for enterovirus and parechovirus surveillance in Scotland, 2010 to 2012. Eurosurveillance 2014; 19: 20772.

41 Ram D, Manor Y, Gozlan Y, et al. Hepatitis E Virus Genotype 3 in Sewage and Genotype 1 in Acute Hepatitis Cases, Israel. Am J Trop Med Hyg 2016; 95: 216–20.

42 Hassine-Zaafrane M, Sdiri-Loulizi K, Kaplon J, et al. Molecular detection of human noroviruses in influent and effluent samples from two biological sewage treatment plants in the region of Monastir, Tunisia. Food Environ Virol 2014; 6: 125–31.

43 Bisseux M, Debroas D, Mirand A, et al. Monitoring of enterovirus diversity in wastewater by ultra-deep sequencing: An effective complementary tool for clinical enterovirus surveillance. Water Res 2020; 169: 115246.

44 Yan T, O’Brien P, Shelton JM, Whelen AC, Pagaling E. Municipal Wastewater as a Microbial Surveillance Platform for Enteric Diseases: A Case Study for Salmonella and Salmonellosis. Environ Sci Technol 2018; 52: 4869–77.

45 Kumazaki M, Usuku S. Nucleotide Correlations Between Rotavirus C Isolates in Clinical Samples from Outbreaks and in Sewage Samples. Food Environ Virol 2015; 7: 269–75.

46 Fioretti JM, Rocha MS, Fumian TM, et al. Occurrence of human sapoviruses in wastewater and stool samples in Rio De Janeiro, Brazil. J Appl Microbiol 2016; 121: 855–62.

47 Hutinel M, Huijbers PMC, Fick J, Åhrén C, Larsson DGJ, Flach C-F. Population-level surveillance of antibiotic resistance in Escherichia coli through sewage analysis. Euro Surveill 2019; 24. DOI:10.2807/1560-7917.ES.2019.24.37.1800497.

48 Ruggeri FM, Bonomo P, Ianiro G, et al. Rotavirus genotypes in sewage treatment plants and in children hospitalized with acute diarrhea in Italy in 2010 and 2011. Appl Environ Microbiol 2015; 81: 241–9.

49 Fioretti JM, Fumian TM, Rocha MS, et al. Surveillance of Noroviruses in Rio De Janeiro, Brazil: Occurrence of New GIV Genotype in Clinical and Wastewater Samples. Food Environ Virol 2018; 10: 1–6.

50 Kazama S, Masago Y, Tohma K, et al. Temporal dynamics of norovirus determined through monitoring of municipal wastewater by pyrosequencing and virological surveillance of gastroenteritis cases. Water Research 2016; 92: 244–53.

51 Lodder WJ, Buisman AM, Rutjes SA, Heijne JC, Teunis PF, de Roda Husman AM. Feasibility of quantitative environmental surveillance in poliovirus eradication strategies. Appl Environ Microbiol 2012; 78: 3800–5.

52 McCall C, Wu H, Miyani B, Xagoraraki I. Identification of multiple potential viral diseases in a large urban center using wastewater surveillance. Water Research 2020; 184: 116160.

53 Bisseux M, Colombet J, Mirand A, et al. Monitoring human enteric viruses in wastewater and relevance to infections encountered in the clinical setting: a one-year experiment in central France, 2014 to 2015. Euro Surveill 2018; 23. DOI:10.2807/1560-7917.ES.2018.23.7.17-00237.

54 Esteves-Jaramillo A, Estívariz CF, Peñaranda S, et al. Detection of Vaccine-Derived Polioviruses in Mexico Using Environmental Surveillance. The Journal of Infectious Diseases 2014; 210: S315–23.

55 Nakamura T, Hamasaki M, Yoshitomi H, et al. Environmental Surveillance of Poliovirus in Sewage Water around the Introduction Period for Inactivated Polio Vaccine in Japan. Appl Environ Microbiol 2015; 81: 1859–64.

56 Más Lago P, Gary HE, Pérez LS, et al. Poliovirus detection in wastewater and stools following an immunization campaign in Havana, Cuba. Int J Epidemiol 2003; 32: 772–7.

57 Wahjuhono G, Revolusiana null, Widhiastuti D, et al. Switch from oral to inactivated poliovirus vaccine in Yogyakarta Province, Indonesia: summary of coverage, immunity, and environmental surveillance. J Infect Dis 2014; 210 Suppl 1: S347–352.

58 Fumian TM, Leite JPG, Rose TL, Prado T, Miagostovich MP. One year environmental surveillance of rotavirus specie A (RVA) genotypes in circulation after the introduction of the Rotarix® vaccine in Rio de Janeiro, Brazil. Water Res 2011; 45: 5755–63.

59 Delogu R, Battistone A, Buttinelli G, et al. Poliovirus and Other Enteroviruses from Environmental Surveillance in Italy, 2009–2015. Food Environ Virol 2018; 10: 333–42.

60 de Oliveira Pereira JS, da Silva LR, de Meireles Nunes A, de Souza Oliveira S, da Costa EV, da Silva EE. Environmental Surveillance of Polioviruses in Rio de Janeiro, Brazil, in Support to the Activities of Global Polio Eradication Initiative. Food Environ Virol 2016; 8: 27–33.

61 González MM, Fonseca MC, Rodríguez CA, et al. Environmental Surveillance of Polioviruses in Armenia, Colombia before Trivalent Oral Polio Vaccine Cessation. Viruses 2019; 11. DOI:10.3390/v11090775.

62 Richter J, Bashiardes S, Koptides D, et al. 2005 Poliovirus eradication: poliovirus presence in Cyprus 2 years after. Water Science and Technology 2008; 58: 647–51.

63 Coulliette-Salmond AD, Alleman MM, Wilnique P, et al. Haiti Poliovirus Environmental Surveillance. Am J Trop Med Hyg 2019; 101: 1240–8.

64 Chowdhary R, Dhole TN. Interrupting wild poliovirus transmission using oral poliovirus vaccine: environmental surveillance in high-risks area of India. J Med Virol 2008; 80: 1477– 88.

65 Pellegrinelli L, Bubba L, Primache V, et al. Surveillance of poliomyelitis in Northern Italy: Results of acute flaccid paralysis surveillance and environmental surveillance, 2012-2015. Hum Vaccin Immunother 2017; 13: 332–8.

66 Johnson Muluh T, Hamisu AW, Craig K, et al. Contribution of Environmental Surveillance Toward Interruption of Poliovirus Transmission in Nigeria, 2012–2015. J Infect Dis 2016; 213: S131–5.

67 Deshpande JM, Shetty SJ, Siddiqui ZA. Environmental surveillance system to track wild poliovirus transmission. Appl Environ Microbiol 2003; 69: 2919–27.

68 Manor Y, Shulman LM, Kaliner E, et al. Intensified environmental surveillance supporting the response to wild poliovirus type 1 silent circulation in Israel, 2013. Euro Surveill 2014; 19: 20708.

69 Yoshida H, Horie H, Matsuura K, Miyamura T. Characterisation of vaccine-derived polioviruses isolated from sewage and river water in Japan. Lancet 2000; 356: 1461–3.

70 Shulman LM, Manor Y, Sofer D, et al. Neurovirulent Vaccine-Derived Polioviruses in Sewage from Highly Immune Populations. PLOS ONE 2006; 1: e69.

71 Pavlov DN. Poliovirus vaccine strains in sewage and river water in South Africa. Can J Microbiol 2006; 52: 717–23.

72 Tao Z, Wang H, Xu A, et al. Isolation of a recombinant type 3/type 2 poliovirus with a chimeric capsid VP1 from sewage in Shandong, China. Virus Res 2010; 150: 56–60.

73 Vinjé J, Gregoricus N, Martin J, et al. Isolation and characterization of circulating type 1 vaccine-derived poliovirus from sewage and stream waters in Hispaniola. J Infect Dis 2004; 189: 1168–75.

74 Manor Y, Blomqvist S, Sofer D, et al. Advanced Environmental Surveillance and Molecular Analyses Indicate Separate Importations Rather than Endemic Circulation of Wild Type 1 Poliovirus in Gaza District in 2002. Appl Environ Microbiol 2007; 73: 5954–8.

75 Grabow WO, Botma KL, de Villiers JC, Clay CG, Erasmus B. Assessment of cell culture and polymerase chain reaction procedures for the detection of polioviruses in wastewater. Bull World Health Organ 1999; 77: 973–80.

76 Lizasoain A, Burlandy FM, Victoria M, Tort LFL, da Silva EE, Colina R. An Environmental Surveillance in Uruguay Reveals the Presence of Highly Divergent Types of Human Enterovirus Species C and a High Frequency of Species A and B Types. Food Environ Virol 2018; 10: 343–52.

77 Wong MVM, Hashsham SA, Gulari E, Rouillard J-M, Aw TG, Rose JB. Detection and characterization of human pathogenic viruses circulating in community wastewater using multi target microarrays and polymerase chain reaction. J Water Health 2013; 11: 659–70.

78 Pellegrinelli L, Binda S, Chiaramonte I, et al. Detection and distribution of culturable Human Enteroviruses through environmental surveillance in Milan, Italy. J Appl Microbiol 2013; 115: 1231–9.

79 Majumdar M, Martin J. Detection by Direct Next Generation Sequencing Analysis of Emerging Enterovirus D68 and C109 Strains in an Environmental Sample From Scotland. Front Microbiol 2018; 9. DOI:10.3389/fmicb.2018.01956.

80 Cesari C, Colucci ME, Veronesi L, et al. Detection of enteroviruses from urban sewage in Parma. Acta Biomed 2010; 81: 40–6.

81 Ozawa H, Yoshida H, Usuku S. Environmental Surveillance Can Dynamically Track Ecological Changes in Enteroviruses. Appl Environ Microbiol 2019; 85. DOI:10.1128/AEM.01604-19.

82 Farías AA, Mojsiejczuk LN, Pisano MB, et al. Environmental Surveillance of Enteroviruses in Central Argentina: First Detection and Evolutionary Analyses of E14. Food Environ Virol 2018; 10: 121–6.

83 Wieczorek M, Ciąćka A, Witek A, Kuryk Ł, Żuk-Wasek A. Environmental Surveillance of Non-polio Enteroviruses in Poland, 2011. Food Environ Virol 2015; 7: 224–31.

84 Majumdar M, Sharif S, Klapsa D, et al. Environmental Surveillance Reveals Complex Enterovirus Circulation Patterns in Human Populations. Open Forum Infectious Diseases 2018; 5. DOI:10.1093/ofid/ofy250.

85 Pellegrinelli L, Galli C, Binda S, et al. Molecular Characterization and Phylogenetic Analysis of Enteroviruses and Hepatitis A Viruses in Sewage Samples, Northern Italy, 2016. Food Environ Virol 2019; 11: 393–9.

86 Zheng H, Lu J, Zhang Y, et al. Prevalence of nonpolio enteroviruses in the sewage of Guangzhou city, China, from 2009 to 2012. Appl Environ Microbiol 2013; 79: 7679–83.

87 Farkas K, Cooper DM, McDonald JE, Malham SK, de Rougemont A, Jones DL. Seasonal and spatial dynamics of enteric viruses in wastewater and in riverine and estuarine receiving waters. Science of The Total Environment 2018; 634: 1174–83.

88 Grøndahl-Rosado RC, Yarovitsyna E, Trettenes E, Myrmel M, Robertson LJ. A One Year Study on the Concentrations of Norovirus and Enteric Adenoviruses in Wastewater and A Surface Drinking Water Source in Norway. Food Environ Virol 2014; 6: 232–45.

89 O’Brien E, Nakyazze J, Wu H, et al. Viral diversity and abundance in polluted waters in Kampala, Uganda. Water Res 2017; 127: 41–9.

90 Aw TG, Gin KY-H. Environmental surveillance and molecular characterization of human enteric viruses in tropical urban wastewaters. Journal of Applied Microbiology 2010; 109: 716–30.

91 Farías AA, Mojsiejczuk LN, Flores FS, et al. Environmental Surveillance of Human Enteroviruses in Córdoba City, Argentina: Prevalence and Detection of Serotypes from 2009 to 2014. Food Environ Virol 2019; 11: 198–203.

92 Garcia L a. T, Viancelli A, Rigotto C, et al. Surveillance of human and swine adenovirus, human norovirus and swine circovirus in water samples in Santa Catarina, Brazil. J Water Health 2012; 10: 445–52.

93 Motayo BO, Adeniji AJ, Faneye AO. First molecular detection and VP7 (G) genotyping in group A rotavirus by semi-nested RT-PCR from sewage in Nigeria. Rev Inst Med Trop Sao Paulo 2016; 58. DOI:10.1590/S1678-9946201658074.

94 Kargar M, Javdani N, Najafi A, Tahamtan Y. First molecular detection of group A rotavirus in urban and hospital sewage systems by nested-RT PCR in Shiraz, Iran. J Environ Health Sci Eng 2013; 11: 4.

95 Li D, Gu AZ, Zeng S-Y, Yang W, He M, Shi H-C. Monitoring and evaluation of infectious rotaviruses in various wastewater effluents and receiving waters revealed correlation and seasonal pattern of occurrences. J Appl Microbiol 2011; 110: 1129–37.

96 Barril PA, Fumian TM, Prez VE, et al. Rotavirus seasonality in urban sewage from Argentina: effect of meteorological variables on the viral load and the genetic diversity. Environ Res 2015; 138: 409–15.

97 Kiulia NM, Netshikweta R, Page NA, et al. The detection of enteric viruses in selected urban and rural river water and sewage in Kenya, with special reference to rotaviruses. J Appl Microbiol 2010; 109: 818–28.

98 La Rosa G, Iaconelli M, Pourshaban M, Muscillo M. Detection and molecular characterization of noroviruses from five sewage treatment plants in central Italy. Water Research 2010; 44: 1777–84.

99 Mabasa VV, Meno KD, Taylor MB, Mans J. Environmental Surveillance for Noroviruses in Selected South African Wastewaters 2015-2016: Emergence of the Novel GII.17. Food Environ Virol 2018; 10: 16–28.

100 Tao Z, Xu M, Lin X, et al. Environmental Surveillance of Genogroup I and II Noroviruses in Shandong Province, China in 2013. Sci Rep 2015; 5: 17444.

101 Zhou N, Lin X, Wang S, et al. Environmental Surveillance for Human Astrovirus in Shandong Province, China in 2013. Sci Rep 2014; 4: 7539.

102 Zhou N, Lin X, Wang S, et al. Molecular characterization of classic human astrovirus in eastern China, as revealed by environmental sewage surveillance. J Appl Microbiol 2016; 120: 1436–44.

103 Torres C, Barrios ME, Cammarata RV, et al. High diversity of human polyomaviruses in environmental and clinical samples in Argentina: Detection of JC, BK, Merkel-cell, Malawi, and human 6 and 7 polyomaviruses. Sci Total Environ 2016; 542: 192–202.

104 Bonanno Ferraro G, Mancini P, Veneri C, et al. Evidence of Saffold virus circulation in Italy provided through environmental surveillance. Letters in Applied Microbiology 2020; 70: 102–8.

105 Iaconelli M, Bonanno Ferraro G, Mancini P, et al. Nine-Year Nationwide Environmental Surveillance of Hepatitis E Virus in Urban Wastewaters in Italy (2011-2019). Int J Environ Res Public Health 2020; 17. DOI:10.3390/ijerph17062059.

106 Lun JH, Crosbie ND, White PA. Genetic diversity and quantification of human mastadenoviruses in wastewater from Sydney and Melbourne, Australia. Sci Total Environ 2019; 675: 305–12.

107 Shaheen MNF, Abd El-Daim SE, Ahmed NI, Elmahdy EM. Environmental monitoring of Aichi virus and human bocavirus in samples from wastewater treatment plant, drain, and River Nile in Egypt. Journal of water and health 2020; 18: 30–7.

108 Martins FDC, Ladeia WA, Toledo R dos S, Garcia JL, Navarro IT, Freire RL. Surveillance of Giardia and Cryptosporidium in sewage from an urban area in Brazil. Rev Bras Parasitol Vet 2019; 28: 291–7.

109 Ajonina C, Buzie C, Otterpohl R. The detection of Giardia cysts in a large-scale wastewater treatment plant in Hamburg, Germany. J Toxicol Environ Health A 2013; 76: 509–14.

110 Lamba M, Gupta S, Shukla R, Graham DW, Sreekrishnan TR, Ahammad SZ. Carbapenem resistance exposures via wastewaters across New Delhi. Environ Int 2018; 119: 302–8.

111 Yao Y, Lazaro-Perona F, Falgenhauer L, et al. Insights into a Novel blaKPC-2 -Encoding IncP-6 Plasmid Reveal Carbapenem-Resistance Circulation in Several Enterobacteriaceae Species from Wastewater and a Hospital Source in Spain. Front Microbiol 2017; 8. DOI:10.3389/fmicb.2017.01143.

112 Yang K, Pagaling E, Yan T. Estimating the Prevalence of Potential Enteropathogenic Escherichia coli and Intimin Gene Diversity in a Human Community by Monitoring Sanitary Sewage. Appl Environ Microbiol 2014; 80: 119–27.

113 Harris-Lovett S, Nelson KL, Beamer P, et al. Wastewater Surveillance for SARS-CoV-2 on College Campuses: Initial Efforts, Lessons Learned, and Research Needs. International Journal of Environmental Research and Public Health 2021; 18: 4455.

114 Global Water Pathogen Project. https://www.waterpathogens.org/ (accessed June 10, 2021).

115 Adalja AA, Watson M, Toner ES, Cicero A, Inglesby TV. Characteristics of Microbes Most Likely to Cause Pandemics and Global Catastrophes. In: Inglesby TV, Adalja AA, eds. Global Catastrophic Biological Risks. Cham: Springer International Publishing, 2019: 1–20.

116 Casadevall A, Pirofski LA. Host-pathogen interactions: redefining the basic concepts of virulence and pathogenicity. Infection and immunity 1999; 67: 3703–13.

117 Wilder-Smith A, Osman S. Public health emergencies of international concern: a historic overview. Journal of Travel Medicine 2020; 27. DOI:10.1093/jtm/taaa227.

118 Larsen DA, Wigginton KR. Tracking COVID-19 with wastewater. Nat Biotechnol 2020; published online Sept 21. DOI:10.1038/s41587-020-0690-1.

119 Ansari SA, Farrah SR, Chaudhry GR. Presence of human immunodeficiency virus nucleic acids in wastewater and their detection by polymerase chain reaction. Appl Environ Microbiol 1992; 58: 3984–90.

120 Jensen KE. Presence and destruction of tubercle bacilli in sewage. Bull World Health Organ 1954; 10: 171–9.

121 Chandra K, Arora VK. Tuberculosis and other chronic morbidity profile of sewage workers of Delhi. Indian Journal of Tuberculosis 2019; 66: 144–9.

122 Chau K, Barker L, Budgell E, et al. Systematic Review of Wastewater Surveillance of Antimicrobial Resistance in Human Populations. 2021; published online June 21. DOI:10.20944/preprints202010.0267.v2.

123 Jirků M, Pomajbíková K, Petrželková KJ, Hůzová Z, Modrý D, Lukeš J. Detection of Plasmodium spp. in Human Feces. Emerg Infect Dis 2012; 18: 634–6.

124 Heijnen L, Medema G. Surveillance of influenza A and the pandemic influenza A (H1N1) 2009 in sewage and surface water in the Netherlands. J Water Health 2011; 9: 434–42.

125 Furuse Y. Analysis of research intensity on infectious disease by disease burden reveals which infectious diseases are neglected by researchers. PNAS 2019; 116: 478–83.

126 Béji-Hamza A, Khélifi-Gharbi H, Hassine-Zaafrane M, et al. Qualitative and Quantitative Assessment of Hepatitis A Virus in Wastewaters in Tunisia. Food Environ Virol 2014; 6: 246–52.

